# Development of machine learning model for predicting hospitalization in the prehospital setting

**DOI:** 10.1101/2021.11.29.21266929

**Authors:** Kenichiro Morisawa, Tadahiro Goto, Shigeki Fujitani

**Affiliations:** Department of Emergency and Critical Care Medicine, St. Marianna University School of Medicine, 2-16-1 Sugao, Miyamae, 216-8511, Kawasaki, Kanagawa, Japan; TXP Medical Co., Ltd. 7-3-1, Hongo, Bunkyo, Tokyo, 113-8485, Japan

## Abstract

**Background:** Studies have developed models for predicting patient outcomes for successful risk stratification in the prehospital setting. However, these models generally require many predictors to achieve high prediction ability, resulting in a bar for implementing models in the real clinical setting.

**Objective:** We aimed to develop a simple and implementable machine learning model using automatically-collected data (age, sex, vital signs) to predict patient outcomes during transportation in comparison with National Early Warning Score (NEWS).

**Methods:** This is a retrospective cohort study using data from the ED of three tertiary care hospitals in Japan from April 2017 to March 2020. We included adult patients (aged ≥18 years) who were transported to the ED of participating hospitals. We excluded patients with trauma/injury, cardiac arrest, transferred from other hospitals, patients with missing vital signs data, or having data of obvious outliers. The predictors were patient age, sex, mental status evaluated with Japan Coma Scale, systolic blood pressure, diastolic blood pressure, pulse rate, respiratory rate, and oxygen saturation. The primary outcome was hospitalization. We developed a model using XGBoost.

**Results:** During the study period, 3528 visits transported by emergency medical services were eligible. The median NEWS was 4.0, and 2081 patients were hospitalized. The discrimination ability of the newly developed model was 0.70 (95%CI 0.67-0.73), which was better than those of NEWS 0.64 (95%CI 0.61-0.68). The newly developed model’s performance measures (e.g., sensitivity, specificity) were comparable with NEWS.

**Conclusions:** Our newly developed machine learning model using routinely available data has moderate prediction ability and was better than NEWS.

## INTRODUCTION

In 2020, approximately 6 million patients were transported to the emergency department (ED) by emergency medical service (EMS) in Japan.^1^ Given increasing transportations over decades, early risk stratification of patients in the prehospital setting can improve patient outcomes and enhance efficient resource utilization in the context of integrated community health care.^2^

To address this concern, studies have developed models for predicting patient outcomes for successful risk stratification in the prehospital setting.^2-6^ For example, a multicenter study of 4950 trauma patients developed a model to predict severe injury defined as an Injury Severity Score greater than 15, and the model had a high discrimination ability (C statistic of 0.823).^3^ Another prognostic study using machine learning reported that a model using ≥1000 predictors has good discrimination ability.^5^ However, these models, including machine learning models, generally require many predictors to achieve high prediction ability—this becomes a bar to implement prediction models in the real clinical setting.

A recent study reported the benefit of National Early Warning Score (NEWS) in the prehospital setting.^4,6^ While the NEWS initially used to predict illness severity and deterioration in a hospital setting, its availability in the prehospital setting to predict hospitalization is clinically validated.^4^ Although NEWS is a simple and implementable scoring tool, the calculation poses paramedics further burden.

In this context, we aimed to develop a simple and implementable machine learning model using automatically-collected data (age, sex, vital signs) to predict patient outcomes during transportation. We hypothesized that our machine learning model using routinely available data has better prediction ability compared with NEWS.

## METHODS

### Study design and setting

We retrospectively analyzed data from the ED of three tertiary care hospitals (Hitachi General Hospital, Japanese Red Cross Society Kyoto Daiichi Hospital, Utsunomiya-Saiseikai Hospital) in Japan from April 1, 2017 to March 31, 2020. The EDs of participating hospitals are staffed by emergency attending physicians and have affiliations with transitional and emergency medicine (EM) residency training programs. The prehospital data and medical charts are structured through an electronic medical record system (NEXT Stage ER system, TXP Medical, Co., Ltd. Tokyo), which supports healthcare workers input clinical information as structured data.^7^ The prehospital data were recorded by paramedics. The study protocol was approved by the Ethics Committee of TXP Medical Co., Ltd., and the requirement for informed consent was waived due to the retrospective nature of the study.

### Study participants

We included adult patients (aged ≥18 years) transported to the ED of participating hospitals. We excluded patients with trauma/injury, cardiac arrest, transferred from other hospitals, patients with missing vital signs data, or having data of obvious outliers.

### Candidate predictors and outcome

The predictors for machine learning models were chosen from routinely available data in the prehospital setting. Specifically, the predictors were patient age, sex, mental status evaluated with Japan Coma Scale, systolic blood pressure (mmHg), diastolic blood pressure (mmHg), pulse rate (/min), respiratory rate (/min), and oxygen saturation (%). The primary outcome was hospitalization.

### Statistical analysis

First, we used summary statistics to delineate the characteristics of patients transported by emergency medical services (EMS).

Second, we divided the data into the training set (70% random sample) and the test set (the remaining 30%). In the training set, we developed the machine learning model using gradient-boosted decision tree with *xgboost* package. Gradient-boosted decision tree is an ensemble approach—an additive model of decision trees estimated by gradient descent, which does not require computational power and is easy to deploy. For the gradient-boosted tree model, we have used a grid search strategy to identify the best combination of hyperparameters by using the *ranger* and *caret* packages. In the gradient-boosted tree model, we used 10-fold cross-validation to measure the prediction error with a smaller variance than that from a single train-test set split.

In the test set (30% random sample), we measured the prediction performance of each model by computing C statistics (i.e., the area under the receiver operating characteristic [ROC] curve) and prospective prediction results (e.g., sensitivity, specificity). We compared the discrimination ability using DeLong’s test.

All analyses were conducted using R version 3.6.1 (R Foundation, Vienna, Austria).^17^ A P-value of <0.05 was considered statistically significant.

## RESULTS

From April 1, 2017 through March 31, 2020, there were 25,549 ED visits transported by EMS. Of these, we excluded 2808 children or patients with unknown age, 7223 visits for trauma, follow-up visits, or transportation from other hospitals, 11,990 visits without the information on NEWS, and the remaining 3528 ED visits transported by EMS were eligible for the analysis.

Patient characteristics are shown in **Table 1**. Overall, the median age was 75 years, and 45.6% were female. The most frequent chief complaint was altered mental status, followed by dyspnea, abdominal pain, and fever. Approximately two third of patients were clear mental status. Other vital signs were almost within normal ranges. The median NEWS was 4.0, and 2081 patients were hospitalized.

**Table 1.** Patient characteristics and outcomes in 3528 emergency department visits transported by emergency medical services.

| Variables | n=3528 |
| --- | --- |
| Age (year) | 75.0 (61.0, 83.0) |
| Female | 1608 (45.6%) |
| Frequent chief complaints |  |
| Altered mental status | 357 (10.1%) |
| Dyspnea | 348 (9.9%) |
| Abdominal pain | 278 (7.9%) |
| Fever | 268 (7.6%) |
| Chest pain | 202 (5.7%) |
| Vital signs |  |
| Japan Coma Scale |  |
| 0 (alert) | 2273 (64.4%) |
| 1-3 | 896 (25.4%) |
| 10-30 | 203 (5.8%) |
| 100-300 | 156 (4.4%) |
| Systolic blood pressure (mmHg) | 143.0 (120.0, 168.0) |
| Diastolic blood pressure (mmHg) | 83.0 (70.0, 96.0) |
| Pulse pressure (/min) | 86.0 (73.0, 102.0) |
| Body temperature (°C) | 36.7 (36.3, 37.2) |
| Respiratory rate (/min) | 20.0 (18.0, 24.0) |
| Oxygen saturation median (%) | 97.0 (96.0, 99.0) |
| National Early Warning Score | 4.0 (1.0, 6.0) |
| Hospitalization | 2081 (59.0%) |

Data were presented as median with interquartile range otherwise specified.

The prediction ability of the newly developed model and NEWS are shown in **Table 2** and **Figure 1**. The discrimination ability of the newly developed model was 0.70 (95%CI 0.67-0.73), with a sensitivity of 0.60, specificity of 0.67, positive predictive value of 0.71, and negative predictive value of 0.55. As for NEWS, the discrimination ability was 0.64 (95%CI 0.61-0.68), with a sensitivity of 0.59, specificity of 0.66, positive predictive value of 0.70, and negative predictive value of 0.54. The discrimination ability of newly developed model was better than those of NEWS (0.70 vs. 0.64; P<0.001)

**Table 2.** Prediction ability of the developed machine learning model and National Early Warning Score.

| Variables | AUROC | Sensitivity | Specificity | PPV | NPV |
| --- | --- | --- | --- | --- | --- |
| Gradient-boosted decision tree | 0.70 (0.67-0.73) | 0.60 | 0.67 | 0.71 | 0.55 |
| NEWS | 0.64 (0.61-0.68) | 0.59 | 0.66 | 0.70 | 0.54 |

Abbreviations: AUROC, area under the receiver operating characteristic; PPV, positive predictive value; NPV, negative predictive value; NEWS, National Early Warning Score

**Figure 1.**
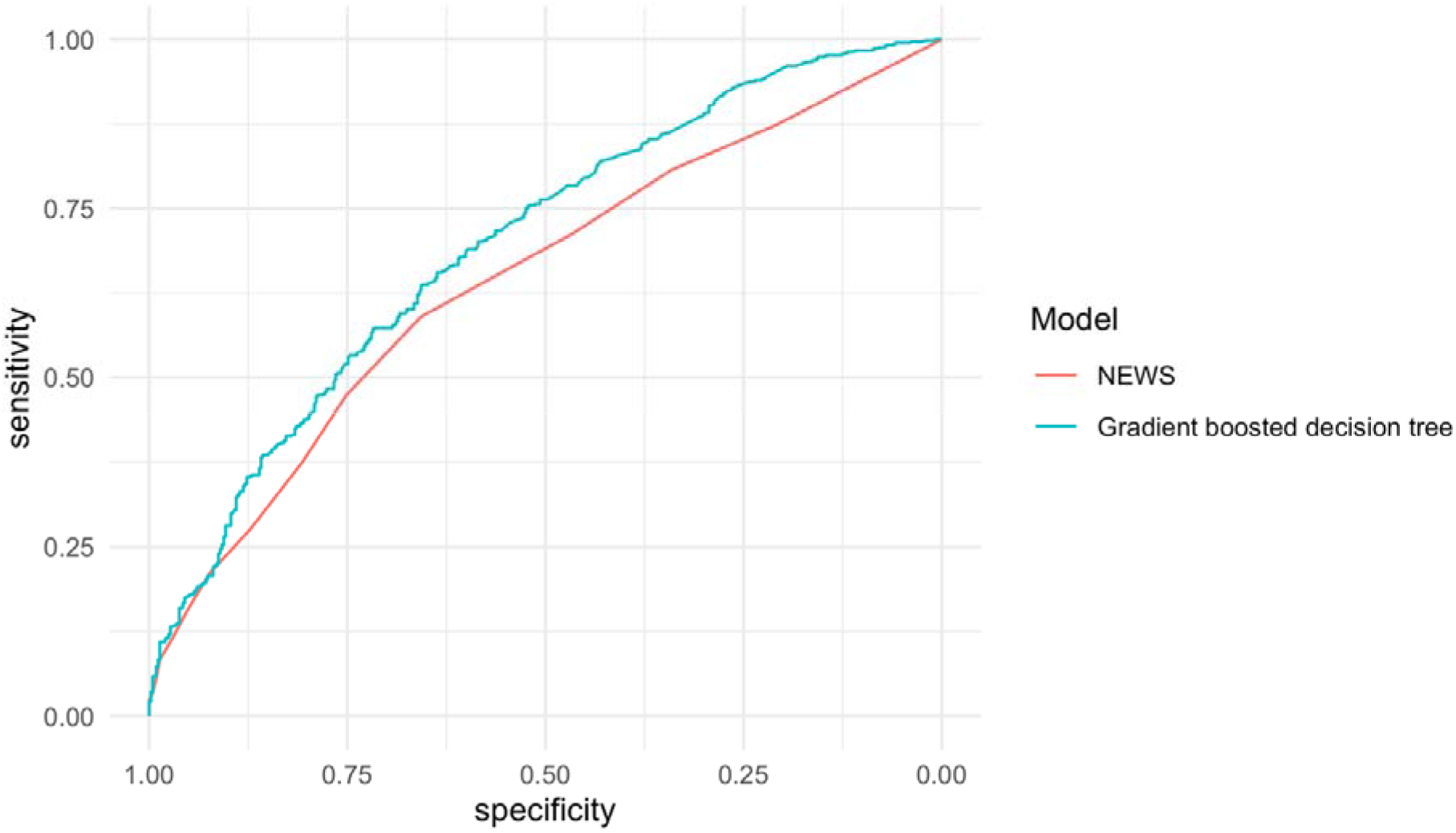
Prediction Ability of the Machine Learning Model and National Early Warning Score.

## DISCUSSION

Our newly developed machine-learning model using the data of 3528 patients transported by EMS has a higher prediction ability compared with NEWS. While we used only routinely available data (age, sex, and vital signs), the model had the moderate prognostic ability to predict hospitalization in the prehospital setting.

A previous study reported that the performance of EMS staffs’ ability to predict patient outcomes (hospitalization) was C statistic of 0.60.^8^ Although direct comparison should not be allowed, the prediction ability of our model might be comparable with paramedic’s evaluation. In addition, since we used only objective measurements, the results may not be affected by paramedics’ experiences.

The strength of the current study is easy-to-implement with routinely available data. Recent technology enables us to use a smartphone app or monitor-implemented scoring systems in the prehospital setting.^9^ Once this machine learning model is implemented in the monitoring system, paramedics can evaluate patient severity automatically just by attaching a monitor to the patient and entering the information on age, gender, and consciousness level. In addition, because we mainly used vital signs, further sophisticated models can be developed based on the current model (e.g., using longitudinal data).

This study has several potential limitations. First, due to the nature of retrospective design, excluding patients with missing data in vital signs may cause biased estimation. Second, a relatively limited sample size to develop a machine learning model can calm the advantages of machine learning, which can address non-linear relationships. Lastly, the model requires further external validation in the different settings.

## CONCLUSIONS

Our newly developed machine learning model has moderate prediction ability and was better than NEWS. Because the developed model requires routinely available data and minimum computational resources, our model is implementable in real prehospital settings.

## Data Availability

All data produced in the present study are available upon reasonable request to the authors

